# Leveraging Pretrained Vision Transformers for Automated Cancer Diagnosis in Optical Coherence Tomography Images

**DOI:** 10.1101/2024.09.26.24314445

**Authors:** Soumyajit Ray, Cheng-Yu Lee, Hyeon-Cheol Park, David W. Nauen, Chetan Bettegowda, Xingde Li, Rama Chellappa

## Abstract

This study presents a novel approach to brain cancer detection based on Optical Coherence Tomography (OCT) images and advanced machine learning techniques. The research addresses the critical need for accurate, real-time differentiation between cancerous and noncancerous brain tissue during neurosurgical procedures. The proposed method combines a pre-trained Vision Transformer (ViT) model, specifically DiNOV2, with a convolutional neural network (CNN) operating on Grey Level Co-occurrence Matrix (GLCM) texture features. This dual-path architecture leverages both the global context capture capabilities of transformers and the local texture analysis strengths of GLCM + CNNs. The dataset comprised OCT images from 11 patients, with 5,831 B-frame slices used for training and validation, and 1,610 slices for testing. The model achieved high accuracy in distinguishing cancerous from noncancerous tissue, with 99.7% ± 0.1% accuracy on the training dataset, 99.4% ± 0.1% on the validation dataset and 94.9% accuracy on the test dataset. This approach demonstrates significant potential for achieving and improving intraoperative decision-making in brain cancer surgeries, offering real-time, high-accuracy tissue classification and surgical guidance.

## Introduction

The accurate detection and maximal excision of cancerous brain tissues is a critical and complex aspect of neurosurgery. Optimal brain cancer resection, which involves excising the maximum amount of malignant tissue while preserving the adjacent noncancerous tissues, is crucial for preventing cancer recurrence and minimizing neurological damage during surgery [1,2]. Distinguishing between noncancerous and cancerous tissue is essential for patient outcomes and quality of life (Jermyn et al., 2015). The surgical intervention of brain cancers has traditionally relied on a combination of imaging techniques like pre-operative Magnetic Resonance Imaging (MRI) and Computed Tomography (CT) scans, supplemented by intra-operative frozen section histopathological examination. These traditional methods, while having made a significant impact, have limitations in terms of providing real-time guidance [1,2]. A surgical microscope is widely used for intra-operative guidance; however, it only provides magnified view of the tissue surface and direct detection of infiltrative tissues is challenging. Optical coherence tomography (OCT), a non-invasive imaging method, has become a powerful tool in this field, providing high-resolution, cross-sectional images of tissue microstructures. OCT’s ability to provide real-time, microscopic-level images can significantly improve intraoperative decision-making.

Recent advancements in OCT technology have shown promising results in differentiating between cancerous and noncancerous brain tissues [3,4]. Studies have demonstrated that optical attenuation of the tissue can be used to distinguish between malignant and benign brain tissue. Optical attenuation is typically lower in cancerous tissue because of the degeneration of myelin sheath in the white matter. This reduction in optical attenuation can be detected in 3D OCT images obtained *in vivo*, using state-of-the art computer parallel processing algorithms. Past work in this area has used texture features to develop a classifier for the OCT images. In our earlier paper [8], we used a combination of texture features combined with a custom designed convolutional neural network (CNN) to extract additional discriminative features from the underlying OCT images. In this work we further extend this by using pretrained neural networks and state of the art transformer models. Transfer learning is a standard practice in computer vision, where a neural network trained on an unrelated dataset can be finetuned using a smaller relevant dataset to generate relevant features for classification. Deep neural networks learn features in a hierarchical manner. The layers of the network typically learn to recognize simple, low-level features (like edges, corners, or color gradients in image processing) that are invariant or equivariant to certain transformations. We also incorporate Vision Transformers (ViTs) which have demonstrated remarkable performance on various image understanding tasks, often surpassing traditional CNNs. The self-attention mechanism built in transformer architecture allows the model to weigh the importance of various parts of the input when processing each element. This is particularly useful to combine local and global image features in the OCT image. The success of these models typically relies on extensive labeled datasets. Self-supervised pretraining aims to address this limitation by enabling models to learn meaningful representations from unlabeled data. In this work we use a pretrained ViT – DinOV2 [14] - to extract relevant features from the OCT images. DinoV2 implements several key innovations that improve upon other self-supervised pretrained vision models. A multi-scale architecture enables processing images at different resolutions, which allows the model to capture both fine-grained details and the global context more effectively. DinoV2 has been trained on a pretraining dataset consisting of 142 million images.

## Dataset

The dataset consists of OCT images of brain tissue obtained from the 11 patients. The average age of the patients was 62.6 years (age range: 33 – 78 years). Of these 11 patients, 7 patients were used in the training set – 4 with cancerous brain tissue and 3 noncancerous. 5,831 B-frame slices were obtained from these patients. Of these 20% were used as the validation dataset and the rest were used for training. The testing set consisted of images from 4 patients – 2 cancerous and 2 non–cancerous patients – for 1,610 slices. In line with our previous work [8], a 100 × 100 grey level co-occurrence matrix (GLCM) was extracted from every B-frame slice. The GLCM is hypothesized to capture discriminative information of the texture features in the OCT image.

As detailed in our previous study [8], we adopt the following preprocessing steps. The B-frame data is logarithmically (base 10) transformed. Approximately 25 representative B-frames using a step size of 80 µm are selected. This is because our OCT system has a transverse resolution of 25 µm, allowing for stable B-frame data within this range, thus sparse sampling ensures a representative dataset [8]. B-frames which show significant fragmentation at the tissue surface are manually excluded. This is a small fraction of the total samples. The remaining frames undergo a 5×5 median filter process to reduce noise. Each B-frame is then divided into seven equal sections, each containing 100 A-lines, spaced 290 µm apart. Within each section, the A-lines are shortened to focus only on the single scattering region, which extends about 200 pixels (or 418 µm in depth) below the sample surface. This process results in 5,831 examples, calculated as 34 samples, with about 25 B-frames per sample and seven slices per B-frame.

## Methods

As illustrated in Fig. 1 (B), top row, slices from noncancerous B-frames display a significant white to black gradient from top to bottom compared to those from cancerous frames (Fig. 1 (B)), indicating that noncancerous white matter more rapidly diminishes the OCT intensity along depth. The grey level co-occurrence matrix (GLCM) quantifies the image texture of each B-frame slice to capture the global texture of the image. The GLCM [10] is a well-established method for quantifying the arrangement of pixel values or the spatial relationship of pixels. It has been widely used in various medical imaging applications, such as OCT, to extract distinctive features [9]. Initially, the highest and lowest values in the training set are used to linearly convert the values in each B-frame slice to a new range that spans from 0 to 99, including both endpoints. Next, a texture map of size 100 × 100 is calculated for each slice using a condition of 15 pixels adjacent to each other at an angle of 0 degrees. These parameters were determined experimentally in our previous study [8]. The row and column indices of the GLCM correspond to pixel intensities, while the matrix elements indicate the frequency of adjacency between the two supplied intensities [11]. Figure 1(A) and (B) bottom row displays characteristic textural attributes seen in the training set. Cancer slices are characterized by concentrated circles, while noncancer slices have oblong and diffused textures. The DinoV2 transformer model plays a significant role in this application. Given its training on extremely large dataset of natural images, this transformer is theorized to extract a diverse range of characteristics from an OCT image. These characteristics encompass several elements of the image, such as textures, forms, and patterns, which could potentially indicate the presence of either noncancerous or malignant tissues. The DiNOV2 ‘small’ model used for this study extracts a single vector of length 384 for each input image (embedding vector).

**Figure 1.**
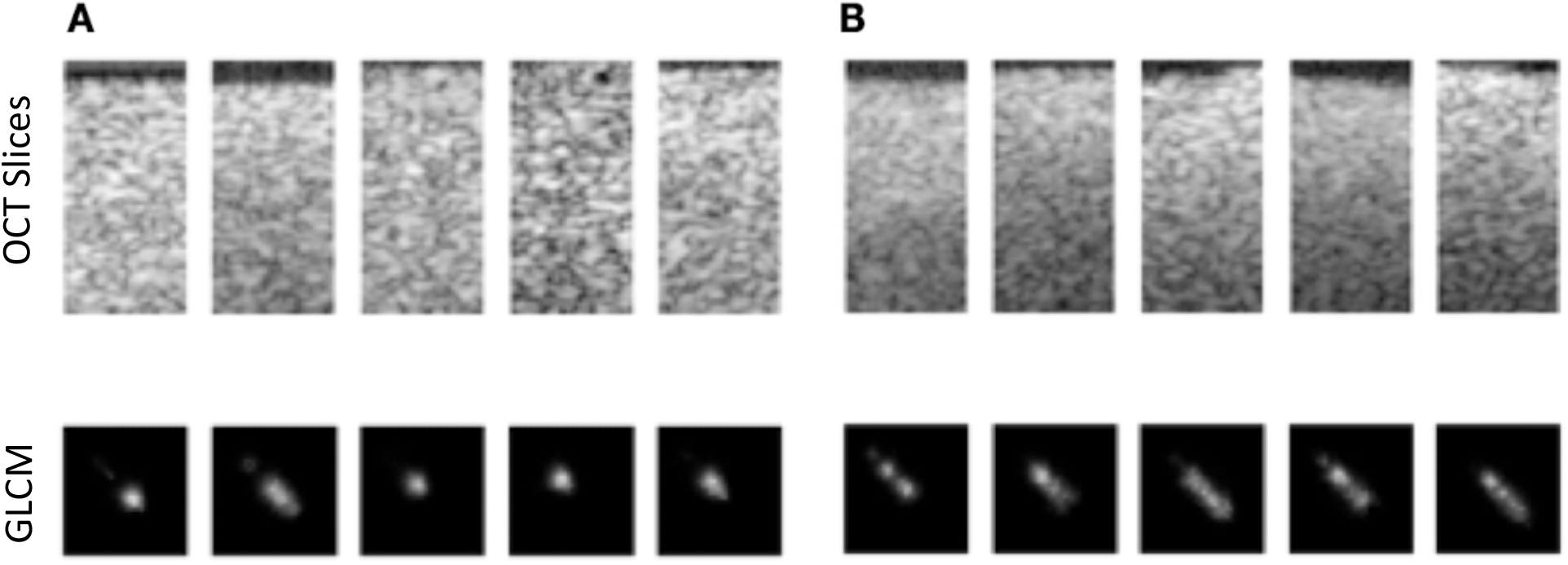
Dataset visualization. (A) Top row are the OCT B-frame slices of dimensions 100 × 200 pixels acquired from the cancerous brain tissues. The images in the bottom row are the corresponding GLCM extracted from the B-frame slices. (B) Top row is the OCT B-frame slices of dimensions 100 × 200 pixels acquired from the non-cancerous tissues. Images in the bottom row are the corresponding GLCM extracted from the B-frame slices. GLCM: Grey Level Co-occurrence Matrix.

The extracted GLCM texture matrix is passed through a pretrained standard ResNet (ResNet18) [12]. This is a convolutional neural which has been pretrained on the ImageNet dataset [13]. The output features of the transformer are combined with the embeddings extracted from the ResNet which processes the GLCM texture matrix. The concatenated embeddings are input to the final classification head. The classification head is a collection of layers in the neural network trained to detect the relevant cancerous tissue patterns. The last layer of the classification head is the softmax activation function. The output of the classification head is a value between 0 and 1 which represents the probability of the input image being cancerous tissue. A value approaching 1 signifies a greater probability of cancer, whereas a value approaching 0 implies a lesser probability. An empirically determined threshold is used to get the final binary classification. Figure 2 depicts the high-level structure of the network.

**Figure 2.**
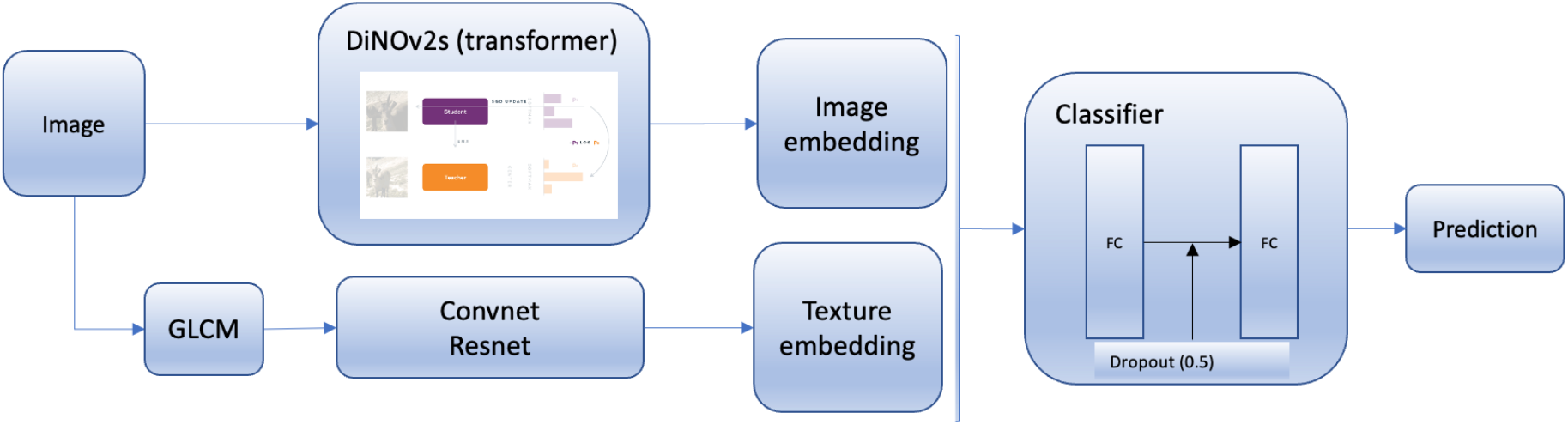
Overall deep learning model architecture. DiNOV2 is used as the backbone for feature extraction from the OCT images of brain tissue. The extracted features/embeddings are combined with the features extracted from the texture processing branch of the model (GLCM + ResNet).

Binary cross entropy [15] is used as the loss function to train this network. It is augmented with balanced class weights regularization to adjust the loss in a way that prevents bias towards the majority class [14]. A batch size of 64 was used during training. The Adam optimizer was used with a learning rate of 1E-4 and an exponential learning rate scheduler to progressively decay the learning to accelerate convergence. Both branches of the model – DiNOV2 transformer and the ResNet, along with the classification head are jointly trained. For the branch of the model which processes the GLCM matrix (ResNet) the entire branch is fine-tuned with the training data. Whereas for the main backbone which processes the OCT image directly the weights of the transformer are kept frozen and only the additional classifier layers are finetuned with the training data. Appropriate data augmentation in the form of random crops and horizontal flipping was used to improve the training process. A dropout rate of 0.5 was implemented for each dense layer to prevent overfitting. To further reduce overfitting, additional regularization in the form of L2 regularization was implemented as weight decay of 0.01 in the Adam optimizer. The hyperparameter settings, such as layer size, feature maps, optimizer, loss, and learning rate, were chosen through empirical research, adhering to general best practices outlined in this section. The network experiments were performed using a NVIDIA RTX A5000 GPU and an AMD Ryzen Threadripper 3960X 24-Core Processor CPU.

## Results

The transformer network combined with the CNN operating on the GLCM texture matrix had an overall accuracy of 99.7% ± 0.1% on the training dataset after 50 epochs of training and an accuracy of 99.4% ± 0.1% on the validation dataset. On the test dataset the model achieves an accuracy of 94.9%.

The training graphs depicted in Fig. 3 show smooth convergence with well-matched training and validation curves. For some epochs, the validation accuracy even surpassed the training accuracy, likely because augmentation and dropout were disabled during validation, simplifying predictions. The model achieved a prediction time of 120 milliseconds per input during inference on a single NVIDIA RTX A5000 GPU. Although this is considerably longer than the 7 milliseconds per B-frame achieved in our previous study with a CNN-based classifier [8], it remains suitable for real-time applications.

**Figure 3.**
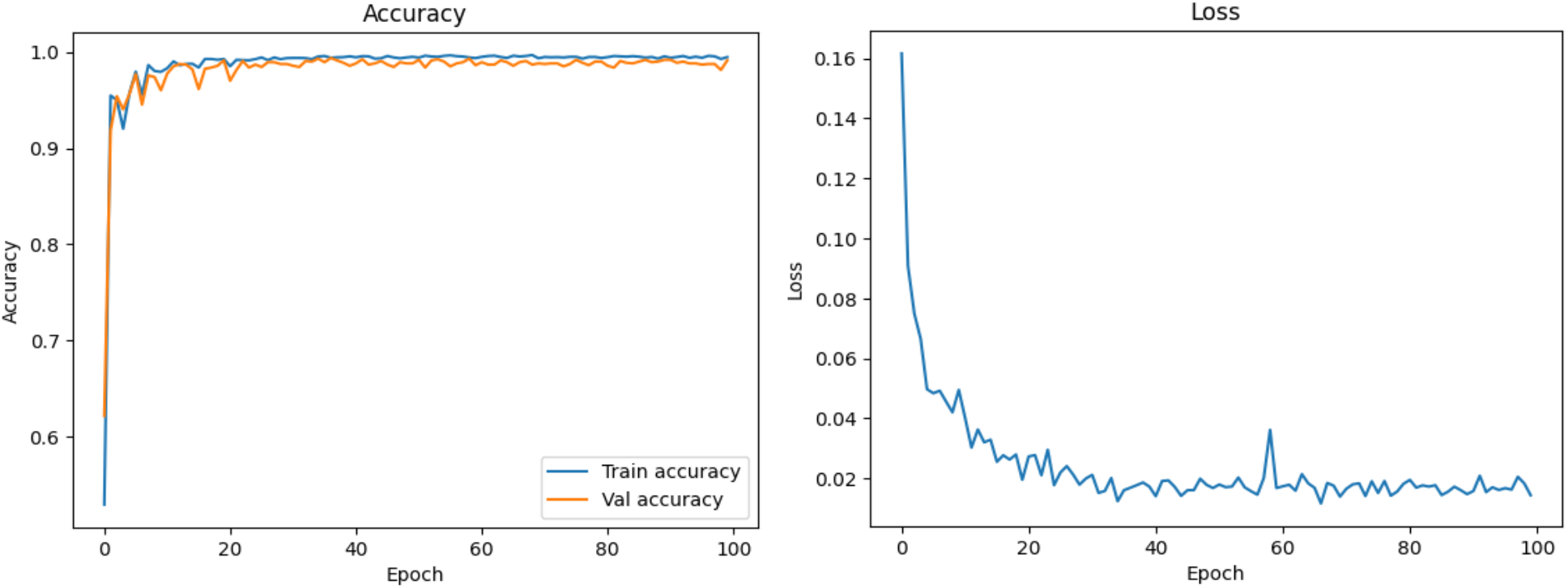
The loss function converges smoothly. Both the training and the validation accuracies are also increasing with some oscillations in the initial training phase. Beyond 40 epochs there is only marginal improvement in the accuracies. The loss also does not decrease much after 40 epochs.

## Discussion

This paper presents a state-of-the-art transformer-based cancer detector capable of accurately detecting cancerous tissue in OCT images in real time. The ensemble model, which integrates a B-frame pretrained transformer model and a texture CNN, is demonstrably superior to any of its component parts in isolation. Given the speed of inference, it is also suitable for applications in real-time cancer detection during the surgical excision of cancerous tissues.

Future directions could potentially include making the model robust to noise in the form of the presence of blood in the tissue and motion artifacts due to movement of the surgeon’s hand or the tissue itself during the imaging process. These sources are noise are more prevalent in *in vivo* imaging. Therefore, it would be beneficial to train the networks on *in vivo* datasets to improve robustness. Training on larger OCT image datasets and further regularizing the network to prevent overfitting should also boost generalization to unseen data. Using large datasets would allow for training a transformer model from scratch, eliminating the need for general-purpose pretrained transformers and resulting in faster inference. Accurately learned feature identification could potentially be used to differentiate various cancer grades (low and high-grade cancers), which will be immensely useful for clinical applications.

## Disclosures

The authors declare no conflicts of interest.

## Data Availability

All data produced in the present study are available upon reasonable request to the authors

## Notes

### Competing Interest Statement

The authors have declared no competing interest.

### Funding Statement

The authors acknowledge the funding support in part by NIH under a grant No. R01CA200399 (Li).

### Author Declarations

Data obtained at the Johns Hopkins Hospital under an approved Institutional Review Board protocol

